## Appendix for "Effects of the COVID-19 pandemic on the mental health of clinically extremely vulnerable children and children living with clinically extremely vulnerable people in Wales: A data linkage study"

**SAIL databank additional information**

**SAIL databank information governance policies and procedures**

The SAIL databank ([www.saildatabank.com](http://www.saildatabank.com)) is an internationally recognised, remotely-accessible, privacy-protecting data safe haven designed to support observational, interventional, and policy-relevant research to improve population health, well-being, and services.^1–6^ SAIL contains anonymised, linkable, routinely collected health, administrative, and social care data for the population of Wales, UK, from multiple sources at individual, household, and ecological levels.^1–6^ Data are anonymised using a split-file process, which has been described in detail elsewhere.^1–3^ Within each dataset, identifiable and non-identifiable data are separated, and identifiable data are sent to a trusted third party (TTP), Digital Health and Care Wales (DHCW; previously known as the NHS Wales Informatics Service). The TTP uniquely matches identities based on name, NHS number, date of birth, and Unique Property Reference Number (UPRN), using the Matching Algorithm for Consistent Results in Anonymised Linkage, which has an accuracy of 99.85%.^1,2^ Individuals and residences are then assigned unique identifiers: for individuals this is called an Anonymised Linking Field (ALF) and for residences a Residential Anonymised Linking Field (RALF). The anonymised and non-identifiable data components are then recombined within SAIL and the linking fields are further encrypted and used to anonymously link between datasets. This enables data from multiple sources, including general practice (GP) data, hospital admissions, outpatient data, and demographic details to be linked at the individual and household level, while preserving anonymity.

**Ethical Approval**

The Information Governance Review Panel (IGRP) is an independent panel of representatives from various government, regulatory, and professional organisations, who review all proposals for SAIL data access to ensure that they are appropriate with respect to Information Governance, and in the public interest.^1^ Data were analysed within the SAIL secure research environment, and appropriate disclosure control procedures were followed to ensure that no personally identifiable data or small numbers (n<5) were removed from the environment. All data within SAIL are treated in accordance with the Data Protection Act 2018 and SAIL complies with the principles of the General Data Protection Regulation (GDPR).

**ICD-10 codes for cancers of the blood or bone marrow**

| **ICD-10 code** | **Description** |
| --- | --- |
| C81 | Hodgkin lymphoma |
| C82 | Follicular lymphoma |
| C83 | Non-follicular lymphoma |
| C84 | Mature T/NK-cell lymphomas |
| C85 | Other and unspecified types of non-Hodgkin lymphoma |
| C86 | Other specified types of T/NK-cell lymphoma |
| C88 | Malignant immunoproliferative diseases |
| C90 | Multiple myeloma and malignant plasma cell neoplasms |
| C91 | Lymphoid leukaemia |
| C92 | Myeloid leukaemia |
| C93 | Monocytic leukaemia |
| C94 | Other leukaemias of specified cell type |
| C95 | Leukaemia of unspecified cell type |
| C96 | Other and unspecified malignant neoplasms of lymphoid, haematopoietic and related tissue |
| D470 | Histiocytic and mast cell tumours of uncertain and unknown behaviour |
| D475 | Chronic eosinophilic leukaemia [hypereosinophilic syndrome] |
| D477 | Other specified neoplasms of uncertain or unknown behaviour of lymphoid, haematopoietic and related tissue |
| D479 | Neoplasm of uncertain or unknown behaviour of lymphoid, haematopoietic and related tissue, unspecified |
| D595 | Paroxysmal nocturnal haemoglobinuria [Marchiafava-Micheli] |
| D71X | Functional disorders of polymorphonuclear neutrophils |
| D730 | Hyposplenism |
| D760 | Langerhans' cell histiocytosis, not elsewhere classified |
| D761 | Haemophagocytic lymphohistiocytosis |
| D898 | Other specified disorders involving the immune mechanism, not elsewhere classified |
| D899 | Disorder involving the immune mechanism, unspecified |
| L412 | Lymphomatoid papulosis |
| P615 | Transient neonatal neutropenia |

**ICD-10 codes for respiratory illnesses**

| **ICD-10 code** | **Description** |
| --- | --- |
| E84 | Cystic Fibrosis |
| J84 | Other interstitial pulmonary diseases |
| J620 | Pneumoconiosis due to talc dust |
| J630 | Aluminosis (of lung) |
| J631 | Bauxite fibrosis (of lung) |
| J633 | Graphite fibrosis (of lung) |
| J634 | Siderosis |
| J635 | Stannosis |
| J660 | Byssinosis |
| J661 | Flax-dresser disease |
| J662 | Cannabinosis |
| J668 | Airway disease due to other specific organic dusts |
| J670 | Farmer lung |
| J671 | Bagassosis |
| J678 | Hypersensitivity pneumonitis due to other organic dusts |
| J684 | Chronic respiratory conditions due to chemicals, gases, fumes and vapours |
| J688 | Other respiratory conditions due to chemicals, gases, fumes and vapours |
| J698 | Pneumonitis due to other solids and liquids |
| J701 | Chronic and other pulmonary manifestations due to radiation |
| J703 | Chronic drug-induced interstitial lung disorders |
| J840 | Alveolar and parietoalveolar conditions |
| J983 | Compensatory emphysema |
| J991 | Respiratory disorders in other diffuse connective tissue disorders |
| M313 | Wegener granulomatosis |
| P250 | Interstitial emphysema originating in the perinatal period |
| Q334 | Congenital bronchiectasis |

**OPCS-4 codes for immunosuppression therapy**

| **OPCS-4 code** | **Description** |
| --- | --- |
| X353 | Active Inflammatory thyroid eye disease patients currently on weekly intravenous steroid infusion treatment (12 weekly injections regime) or immunospressant |
| X374 | Intramuscular Immunotherapy |
| X385 | Subcutaneous Immunotherapy |
| X891 | Monoclonal antibodies Band 1 |
| X892 | Monoclonal antibodies Band 2 |
| X893 | Patients receiving maintenance treatment with rituximab, obinotuzimab or ofatumumab |
| X894 | Somatostatin analogues Band 1 |
| X895 | Allergic emergency drugs Band 1 |
| X961 | Patients previously treated for haematological malignancy requiring IV immunoglobulin replacement |
| X962 | Allergen immunotherapy drugs Band 1 |
| X963 | Poison management drugs Band 1 |

**Pre-COVID-19 study cohorts**

Figure 3 shows a flow diagram of inclusion criteria for the two 2019 study cohorts: children in the general population, and clinically extremely vulnerable children with blood or bone cancer or respiratory illnesses, or receiving immunosuppression therapy.

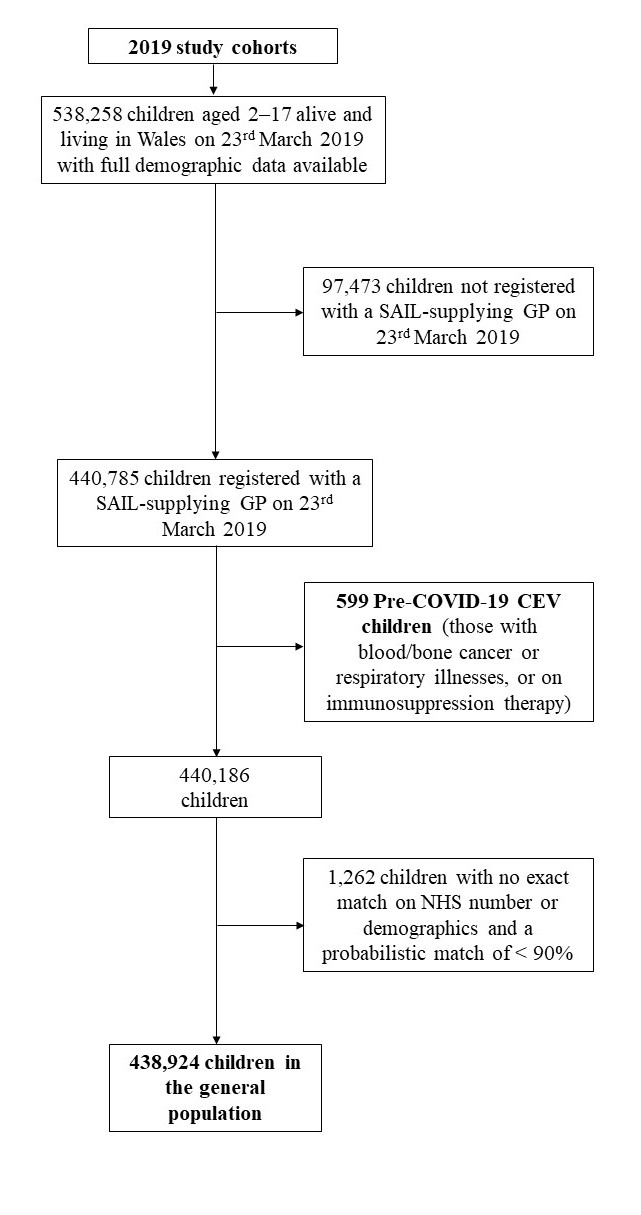

**Figure 3. Flow diagram of the inclusion criteria for the creation of the 2019 study cohorts**

**Sensitivity Analysis**

We undertook a sensitivity analysis to confirm the validity of creating a cohort of pre-COVID-19 CEV children based on just three of the categories included in the COVID-19 Shielded People List (CVSP). We created a cohort of clinically extremely vulnerable (CEV) children who were added to the CVSP in 2020 for the same three reasons only (respiratory illnesses, blood/bone cancer, and immunosuppression therapy) and a cohort of children in the general population (figure 1). Demographic characteristics of the children in each cohort are presented in Table 1. We plotted the Kaplan-Meier survival curve for each cohort and estimated the hazard ratio of having a record for anxiety or depression during the COVID-19 pandemic (March 23^rd^ 2020–January 31^st^ 2021) using Cox regression (figure 2).

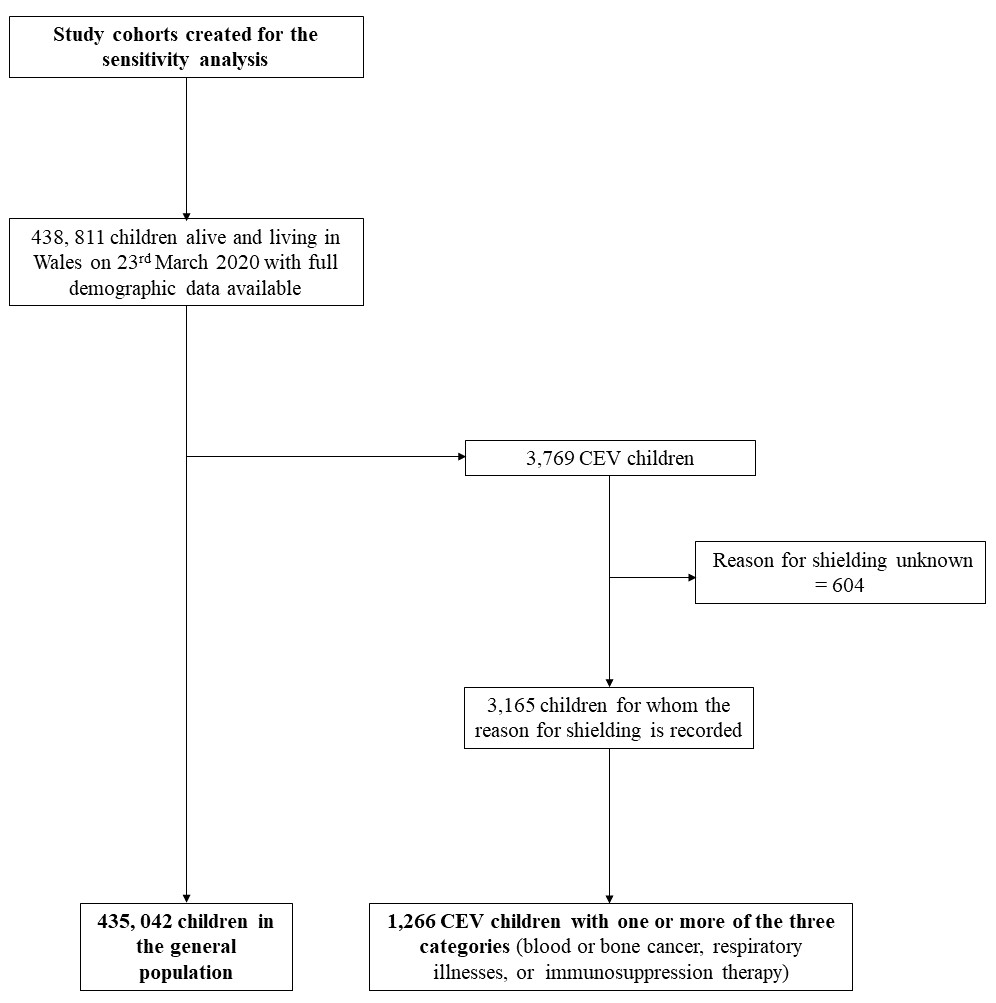

**Figure 1. Flow diagram of the inclusion criteria for the creation of the study cohorts for the sensitivity analysis.**

|  | | General population 2020 | CEV children with one or more of three categories 2020 | Chi^2^ P value |
| --- | --- | --- | --- | --- |
| N | | 435,042 | 1,266 |  |
| **Sex (%)** | Male | 222,558 (51.2) | 712 (56.2) | <0.001 |
|  | Female | 212,484 (48.8) | 554 (43.8) |  |
| **Age group (%)** | 2–7 | 157,691 (36.2) | 385 (30.4) | <0.001 |
|  | 8–12 | 143,988 (33.1) | 396 (31.3) |  |
|  | 13–17 | 133,363 (30.7) | 485 (38.3) |  |
| **Deprivation quintile (WIMD 2019) (%)** | 1 (most deprived) | 111,133 (25.5) | 329 (26.0) | 0.262 |
|  | 2 | 92,037 (21.2) | 257 (20.3) |  |
|  | 3 | 76,717 (17.6) | 210 (16.6) |  |
|  | 4 | 73,277 (16.8) | 203 (16.0) |  |
|  | 5 (least deprived) | 81,878 (18.8) | 267 (21.1) |  |
| **Rural/Urban area (%)** | Rural | 116,126 (26.7) | 336 (26.5) | 0.928 |
|  | Urban | 318,916 (73.3) | 930 (73.5) |  |
| **Any history of anxiety or depression** | NO | 415,925 (95.6) | 1,166 (92.1) | <0.001 |
|  | YES | 19,117 (4.4) | 100 (7.9) |  |

**Table 1. Demographic characteristics of children in the general population and clinically extremely vulnerable children added to the COVID-19 Shielded People List for one or more of three reasons (blood or bone cancer, respiratory illnesses, or immunosuppression therapy)**

The results showed a similar pattern to those obtained when analysing the full CEV cohort in that CEV children with one or more of the three categories were at greater risk of having a record for anxiety or depression during the COVID-19 pandemic (March 23^rd^ 2020–January 31^st^ 2021) compared to children in the general population, thereby validating our approach.

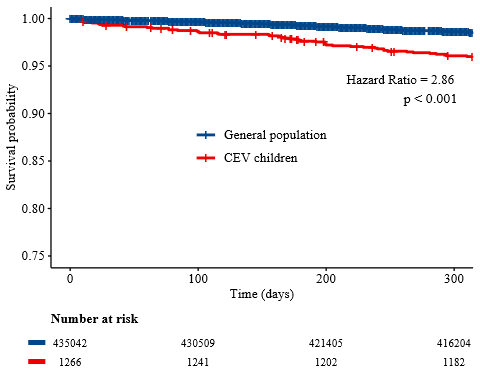

**Figure 2. Kaplan-Meier survival curves for each cohort, showing the time to first record of anxiety or depression during the COVID-19 pandemic**

CEV, clinically extremely vulnerable

**Read v2 and ICD-10 diagnosis codes used to identify records of anxiety or depression in primary or secondary care**

**Read v2 codes for depression diagnosis**

| **Read v2 code** | **Description** |
| --- | --- |
| Eu32. | [X]Depressive episode |
| Eu320 | [X]Mild depressive episode |
| Eu321 | [X]Moderate depressive episode |
| Eu322 | [X]Severe depressive episode without psychotic symptoms |
| Eu324 | [X]Mild depression |
| Eu32y | [X]Other depressive episodes |
| Eu32z | [X]Depressive episode, unspecified |
| Eu33. | [X]Recurrent depressive disorder |
| Eu330 | [X]Recurrent depressive disorder, current episode mild |
| Eu331 | [X]Recurrent depressive disorder, current episode moderate |
| Eu332 | [X]Recurrent depressive disorder, current episode severe without psychotic symptoms |
| Eu334 | [X]Recurrent depressive disorder, currently in remission |
| Eu33y | [X]Other recurrent depressive disorders |
| Eu33z | [X]Recurrent depressive disorder, unspecified |
| Eu341 | [X]Dysthymia |
| E118. | Seasonal affective disorder |
| E135. | Agitated depression |
| E2B.. | Depressive disorder NEC |
| E2B1. | Chronic depression |
| E291. | Prolonged depressive reaction |
| E204. | Neurotic depression reactive type |
| E2B0. | Postviral depression |
| E112. | Single major depressive episode |
| E1120 | Single major depressive episode, unspecified |
| E1121 | Single major depressive episode, mild |
| E1122 | Single major depressive episode, moderate |
| E1123 | Single major depressive episode, severe, without psychosis |
| E1125 | Single major depressive episode, partial or unspecied remission |
| E1126 | Single major depressive episode, in full remission |
| E112z | Single major depressive episode NOS |
| E113. | Recurrent major depressive episode |
| E1130 | Recurrent major depressive episodes, unspecified |
| E1131 | Recurrent major depressive episodes, mild |
| E1132 | Recurrent major depressive episodes, moderate |
| E1133 | Recurrent major depressive episodes, severe, no psychosis |
| E1135 | Recurrent major depressive episodes, partial/unspecified remission |
| E1136 | Recurrent major depressive episodes, in full remission |
| E1137 | Recurrent depression |
| E113z | Recurrent major depressive episode NOS |

**Read v2 codes for depression symptoms**

| **Read v2 code** | **Description** |
| --- | --- |
| 1B17. | Depressed |
| 1B1U. | Symptoms of depression |
| 1BQ.. | Loss of capacity for enjoyment |
| 1BT.. | Depressed mood |
| 1BU.. | Loss of hope for the future |
| 2257 | O/E – depressed |
| 1BP.. | Loss of interest |

**Read v2 codes for antidepressant prescriptions**

| **Read v2 code** | **Description** |
| --- | --- |
| d71.. | Amitriptyline hydrochloride |
| d72.. | Butriptyline - discontinued |
| d73.. | Clomipramine hydrochloride |
| d74.. | Desipramine hydrochloride |
| d75.. | Dosulepin Hydrochloride |
| d76.. | Doxepin |
| d77.. | Imipramine hydrochloride |
| d78.. | Iprindole |
| d79.. | Lofepramine |
| d7a.. | Maprotiline hydrochloride |
| d7b.. | Mianserin hydrochloride |
| d7c.. | Nortriptyline |
| d7d.. | Protriptyline hydrochloride |
| d7e.. | Trazadone hydrochloride |
| d7f.. | Trimipramine |
| d7g.. | Viloxazine hydrochloride |
| d7h.. | Amoxapine |
| d81.. | Phenelzine |
| d83.. | Isocarboxazid |
| d84.. | Tranylcypromine |
| d85.. | Moclobemide |
| d91.. | Compound Antidepressants A-Z |
| da1.. | Flupentixol [Antidepressant] |
| da2.. | Tryptophan |
| da3.. | Fluvoxamine Maleate |
| da4.. | Fluoxetine hydrochloride |
| da5.. | Sertraline hydrochloride |
| da6.. | Paroxetine hydrochloride |
| da7.. | Venlafaxine |
| da9.. | Citalopram |
| daA.. | Reboxetine |
| daB.. | Mirtazapine |
| daC.. | Escitalopram |
| daD.. | Agomelatine |
| gde.. | Duloxetine |
| d911. | Limbitrol 5 capsules - discontinued |
| d912. | Limbitrol 10 capsules - discontinued |
| d913. | Motipress tablets |
| x28CP | Discontinued |
| d914. | Motival tablets discontinued |
| d916. | Triptafen tablets - only one not discontinued? |
| d917. | Triftafen-M tablets - discontinued |
| d8… | Monoamine-oxidase |
| d82.. | Iproniazid |

**Read v2 codes for anxiety diagnosis**

| **Read v2 code** | **Description** |
| --- | --- |
| Eu41. | [X]Other anxiety disorders |
| Eu410 | [X]Panic disorder [episodic paroxysmal anxiety] |
| Eu411 | [X]Generalized anxiety disorder |
| Eu413 | [X]Other mixed anxiety disorders |
| Eu41y | [X]Other specified anxiety disorders |
| Eu41z | [X]Anxiety disorder, unspecified |
| E200. | Anxiety states |
| E2000 | Anxiety state unspecified |
| E2001 | Panic disorder |
| E2002 | Generalised anxiety disorder |
| E2004 | Chronic anxiety |
| E2005 | Recurrent anxiety |
| E200z | Anxiety state NOS |
| E202 | Phobic disorders |
| Eu40. | Phobic anxiety disorder |
| Eu930 | [X]Separation anxiety disorder of childhood * |
| Eu931 | [X]Phobic anxiety disorder of childhood* |
| Eu932 | [X]Social anxiety disorder of childhood* |
| E2D0. | Disturbance of anxiety and fearfulness in childhood and adolescence* |
| E2D00 | Childhood and adolescent overanxiousness disturbance* |
| E2D0z | Disturbance of anxiety and fearfulness in childhood and adolescence NOS* |

**Read v2 codes for anxiety symptoms**

| **Read v2 code** | **Description** |
| --- | --- |
| 1B13. | Anxiousness |
| 2258 | O/E - anxious |
| 1B12. | Nerves, nervousness |
| R2y2. | (D) nervousness |
| 2259 | O/E nervous |
| 225J. | O/E panic attack |
| 1B1V. | C/O panic attack |

**Read v2 codes for anxiety prescriptions – hypnotics**

| **Read v2 code** | **Description** |
| --- | --- |
| d11.. | Chloral hydrate |
| d12.. | Clomethiazole edisylate (hypnotic) |
| d13.. | Dichloralphenazone - discontinued |
| d14.. | Flumtrazepam - discontinued |
| d15.. | Flurazepam |
| d16.. | Loprazolam |
| d17.. | Lormetazepam |
| d18.. | Nitrazepam |
| d1a.. | Temazepam (hynotic) |
| d1b.. | Triazolam - discontinued |
| d1c.. | Triclofos sodium |
| d1d.. | Zopiclone |
| d1f.. | Zolpidem |
| d1g.. | Zaleplon |
| d1h.. | Melatonin |
| d1i.. | Dexmedetomidine |

**Read v2 codes for anxiety prescriptions – hypnotics**

| **Read v2 code** | **Description** |
| --- | --- |
| d21.. | Diazepam |
| d22.. | Alprazolam |
| d23.. | Bromazepam |
| d24.. | Chlordiazepoxide |
| d25.. | Chlormezanone |
| d26.. | Clobazam |
| d27.. | Clorazepate dipotassium |
| d28.. | Hydroxyzine hcl (anxiolytic) |
| d29.. | Ketazolam - discontinued |
| d2a.. | Lorazepam (anxiolytic) |
| d2b.. | Medazepam - discontinued |
| d2c.. | Meprobamate |
| d2d.. | Oxazepam |
| d2e.. | Prazepam - discontinued |
| d2f.. | Buspirone hydrocholoride |
| d2g.. | Flumazenil |

**Read v2 codes for mixed anxiety and depression diagnosis**

| **Read v2 code** | **Description** |
| --- | --- |
| E2003 | Anxiety with depression |
| Eu412 | [X]Mixed anxiety and depressive disorder |

**ICD-10 diagnosis codes for depression**

| **ICD-10 code** | **Description** |
| --- | --- |
| F321 | Moderate depressive episode |
| F322 | Severe depressive episode without psychotic symptoms |
| F328 | Other depressive episodes |
| F329 | Depressive episode, unspecified |
| F33 | Recurrent depressive disorder |
| F330 | Recurrent depressive disorder, current episode mild |
| F331 | Recurrent depressive disorder, current episode moderate |
| F332 | Recurrent depressive disorder, current episode severe without psychotic symptoms |
| F334 | Recurrent depressive disorder, currently in remission |
| F338 | Other recurrent depressive disorders |
| F339 | Recurrent depressive disorder, unspecified |
| F341 | Dysthymia |

**ICD-10 diagnosis codes for anxiety and mixed anxiety and depression**

| **ICD-10 code** | **Description** |
| --- | --- |
| F40 | Phobic anxiety disorders |
| F400 | Agoraphobia |
| F401 | Social phobias |
| F402 | Specific (isolated) phobias |
| F408 | Other phobic anxiety disorders |
| F41 | Other anxiety disorders |
| F410 | Panic disorder (episodic paroxysmal anxiety) |
| F411 | Generalized anxiety disorder |
| F413 | Other mixed anxiety disorders |
| F418 | Other specified anxiety disorders |
| F419 | Anxiety disorder, unspecified |
| F930 | Separation anxiety disorder of childhood |
| F931 | Phobic anxiety disorder of childhood |
| F932 | Social anxiety disorder of childhood |
| F412 | Mixed anxiety and depressive disorder |

**The RECORD statement – checklist of items, extended from the STROBE statement, that should be reported in observational studies using routinely collected health data.**

|  | **Item No.** | **STROBE items** | **Location in manuscript where items are reported** | **RECORD items** | **Location in manuscript where items are reported** |
| --- | --- | --- | --- | --- | --- |
| **Title and abstract** | | | | | |
|  | 1 | (a) Indicate the study’s design with a commonly used term in the title or the abstract (b) Provide in the abstract an informative and balanced summary of what was done and what was found | (a) The study design is indicated as a data linkage study in the title  (b) We have provided an informative and balanced summary in the abstract | RECORD 1.1: The type of data used should be specified in the title or abstract. When possible, the name of the databases used should be included.  RECORD 1.2: If applicable, the geographic region and timeframe within which the study took place should be reported in the title or abstract.  RECORD 1.3: If linkage between databases was conducted for the study, this should be clearly stated in the title or abstract. | 1.1 The type of data used is specified in the abstract. It was not possible to name all of the databases used in the abstract.  1.2 We have reported the geographic region within which the study took place in the title and the abstract, and the timeframe in the abstract  1.3 It is clearly stated in the title and the abstract that linkage between databases was conducted for the study |
| **Introduction** | | | | | |
| Background rationale | 2 | Explain the scientific background and rationale for the investigation being reported | The scientific background and rationale for the investigation is explained in the introduction |  |  |
| Objectives | 3 | State specific objectives, including any prespecified hypotheses | The objectives are stated in the introduction |  |  |
| **Methods** | | | | | |
| Study Design | 4 | Present key elements of study design early in the paper | The study design is reported at the beginning of the methods section, under the subheading “study design and data source” |  |  |
| Setting | 5 | Describe the setting, locations, and relevant dates, including periods of recruitment, exposure, follow-up, and data collection | The setting, locations, and relevant dates are described in the “study population and setting” section |  |  |
| Participants | 6 | *(a) Cohort study* - Give the eligibility criteria, and the sources and methods of selection of participants. Describe methods of follow-up  *Case-control study* - Give the eligibility criteria, and the sources and methods of case ascertainment and control selection. Give the rationale for the choice of cases and controls  *Cross-sectional study* - Give the eligibility criteria, and the sources and methods of selection of participants  *(b) Cohort study* - For matched studies, give matching criteria and number of exposed and unexposed  *Case-control study* - For matched studies, give matching criteria and the number of controls per case | (a) Eligibility criteria, and the sources and methods of selection and follow-up of participants are given in the “study population and setting” section  (b) N/A this was not a matched study | RECORD 6.1: The methods of study population selection (such as codes or algorithms used to identify subjects) should be listed in detail. If this is not possible, an explanation should be provided.  RECORD 6.2: Any validation studies of the codes or algorithms used to select the population should be referenced. If validation was conducted for this study and not published elsewhere, detailed methods and results should be provided.  RECORD 6.3: If the study involved linkage of databases, consider use of a flow diagram or other graphical display to demonstrate the data linkage process, including the number of individuals with linked data at each stage. | 6.1 We have described in detail the methods of study population selection in the “study population and setting” section and the “measures” section  6.2 We have referenced the validation studies of the anxiety and depression codes we have used in our study in the “measures” section  6.3 We have provided a flow diagram with the number of individuals included in each cohort at each stage of linkage (Figure 1) |
| Variables | 7 | Clearly define all outcomes, exposures, predictors, potential confounders, and effect modifiers. Give diagnostic criteria, if applicable. | We have clearly defined all relevant measures in the “measures” section | RECORD 7.1: A complete list of codes and algorithms used to classify exposures, outcomes, confounders, and effect modifiers should be provided. If these cannot be reported, an explanation should be provided. | 7.1 We have provided a list of codes used in the study in the appendix |
| Data sources/ measurement | 8 | For each variable of interest, give sources of data and details of methods of assessment (measurement).  Describe comparability of assessment methods if there is more than one group | We have described all data sources and measures in the “study design and data sources” section and the “measures” section |  |  |
| Bias | 9 | Describe any efforts to address potential sources of bias | We have explained that we used a conservative approach to identify cohort participants in the “study population and setting” section |  |  |
| Study size | 10 | Explain how the study size was arrived at | We have included a flowchart (Figure 1) to describe the creation of each of the study cohorts and explained the process of cohort creation, referred to in the “study population and setting” section |  |  |
| Quantitative variables | 11 | Explain how quantitative variables were handled in the analyses. If applicable, describe which groupings were chosen, and why | Handling of quantitative variables is described in the “measures” section |  |  |
| Statistical methods | 12 | (a) Describe all statistical methods, including those used to control for confounding  (b) Describe any methods used to examine subgroups and interactions  (c) Explain how missing data were addressed  (d) *Cohort study* - If applicable, explain how loss to follow-up was addressed  *Case-control study* - If applicable, explain how matching of cases and controls was addressed  *Cross-sectional study* - If applicable, describe analytical methods taking account of sampling strategy  (e) Describe any sensitivity analyses | (a) Statistical methods are described in the “statistical analysis” section  (b) N/A  (c) N/A  (d) N/A  (e) Sensitivity analyses are described in the appendix |  |  |
| Data access and cleaning methods |  | .. |  | RECORD 12.1: Authors should describe the extent to which the investigators had access to the database population used to create the study population.  RECORD 12.2: Authors should provide information on the data cleaning methods used in the study. | 12.1 We described the extent to which the investigators had access to the database population in the “data access and cleaning methods” section  12.2 We provided information on the data cleaning methods in the “data access and cleaning methods” section |
| Linkage |  | .. |  | RECORD 12.3: State whether the study included person-level, institutional-level, or other data linkage across two or more databases. The methods of linkage and methods of linkage quality evaluation should be provided. | 12.3 The methods of linkage and quality evaluation are detailed in the “study design and data source” section and the “study population and setting” section and the flowchart (Figure 1) |
| **Results** | | | | | |
| Participants | 13 | (a) Report the numbers of individuals at each stage of the study (*e.g.*, numbers potentially eligible, examined for eligibility, confirmed eligible, included in the study, completing follow-up, and analysed)  (b) Give reasons for non-participation at each stage.  (c) Consider use of a flow diagram | (a) Numbers of individuals at each study stage are provided in the flow diagrams (Figure 1)  (b) Reasons for exclusion are provided on the flowchart (Figure 1)  (c) We used a flowchart to illustrate selection of our study cohorts (Figure 1) | RECORD 13.1: Describe in detail the selection of the persons included in the study (*i.e.,* study population selection) including filtering based on data quality, data availability and linkage. The selection of included persons can be described in the text and/or by means of the study flow diagram. | 13.1 We described in detail the selection of the persons included in the study in the “study population and setting” section and Figure 1 |
| Descriptive data | 14 | (a) Give characteristics of study participants (*e.g.*, demographic, clinical, social) and information on exposures and potential confounders  (b) Indicate the number of participants with missing data for each variable of interest  (c) *Cohort study* - summarise follow-up time (*e.g.*, average and total amount) | (a) Characteristics of study participants are given in Table 2  (b) N/A  (c) N/A |  |  |
| Outcome data | 15 | *Cohort study* - Report numbers of outcome events or summary measures over time  *Case-control study* - Report numbers in each exposure category, or summary measures of exposure  *Cross-sectional study* - Report numbers of outcome events or summary measures | Numbers of outcome events are reported in the results section in Table 4 |  |  |
| Main results | 16 | (a) Give unadjusted estimates and, if applicable, confounder-adjusted estimates and their precision (e.g., 95% confidence interval). Make clear which confounders were adjusted for and why they were included  (b) Report category boundaries when continuous variables were categorized  (c) If relevant, consider translating estimates of relative risk into absolute risk for a meaningful time period | (a) We report unadjusted and adjusted estimates in the results section, and have explained why we adjusted for each factor  (b) N/A  (c) N/A |  |  |
| Other analyses | 17 | Report other analyses done—e.g., analyses of subgroups and interactions, and sensitivity analyses | We report on sensitivity analyses in the appendix |  |  |
| **Discussion** | | | | | |
| Key results | 18 | Summarise key results with reference to study objectives | Key results are summarised at the beginning of the discussion |  |  |
| Limitations | 19 | Discuss limitations of the study, taking into account sources of potential bias or imprecision. Discuss both direction and magnitude of any potential bias | Limitations are discussed in the ‘Strengths and limitations’ section including potential underestimation of our cohorts | RECORD 19.1: Discuss the implications of using data that were not created or collected to answer the specific research question(s). Include discussion of misclassification bias, unmeasured confounding, missing data, and changing eligibility over time, as they pertain to the study being reported. | We discussed the limitations of the Shielded Patient List in the ‘Strengths and Limitations’ section |
| Interpretation | 20 | Give a cautious overall interpretation of results considering objectives, limitations, multiplicity of analyses, results from similar studies, and other relevant evidence | We have given a cautious and balanced interpretation of the results with reference to the previous literature in the discussion |  |  |
| Generalisability | 21 | Discuss the generalisability (external validity) of the study results | Discussed in the “strengths and limitations” section |  |  |
| **Other Information** | | | | | |
| Funding | 22 | Give the source of funding and the role of the funders for the present study and, if applicable, for the original study on which the present article is based | We have provided the funding source, and the role of the funders in the “Role of the funders” section |  |  |
| Accessibility of protocol, raw data, and programming code |  | .. |  | RECORD 22.1: Authors should provide information on how to access any supplemental information such as the study protocol, raw data, or programming code. | We have provided information on how to access data within the SAIL databank |

*Reference: Benchimol EI, Smeeth L, Guttmann A, Harron K, Moher D, Petersen I, Sørensen HT, von Elm E, Langan SM, the RECORD Working Committee.  The REporting of studies Conducted using Observational Routinely-collected health Data (RECORD) Statement.  *PLoS Medicine* 2015; in press.

*Checklist is protected under Creative Commons Attribution ([CC BY](http://creativecommons.org/licenses/by/4.0/)) license.

**Demographic characteristics of the 2019 study population**

|  | | **General population 2019** | **CEV children 2019** | Chi^2^ P value |
| --- | --- | --- | --- | --- |
| N | | 438,924 | 599 |  |
| **Sex (%)** | Male | 224,754 (51.2) | 347 (57.9) | <0.001 |
|  | Female | 214,170 (48.8) | 252 (42.1) |  |
| **Age group (%)** | 2–7 | 162,730 (37.1) | 231 (38.6) | 0.412 |
|  | 8–12 | 143,844 (32.8) | 181 (30.2) |  |
|  | 13–17 | 132,350 (30.2) | 187 (31.2) |  |
| **Deprivation quintile (WIMD 2019) (%)** | 1 (most deprived) | 111,381 (25.4) | 134 (22.4) | 0.003 |
|  | 2 | 93,046 (21.2) | 137 (22.9) |  |
|  | 3 | 77,600 (17.7) | 88 (14.7) |  |
|  | 4 | 74,288 (16.9) | 94 (15.7) |  |
|  | 5 (least deprived) | 82,609 (18.8) | 146 (24.4) |  |
| **Rural/Urban area (%)** | Rural | 118,418 (27.0) | 165 (27.5) | 0.790 |
|  | Urban | 320,506 (73.0) | 434 (72.5) |  |
| **Any history of anxiety or depression** | NO | 421,659 (96.1) | 565 (94.3) | 0.037 |
|  | YES | 17,265 (3.9) | 34 (5.7) |  |

**Multivariable analysis of risk factors for having a record of anxiety or depression during the COVID-19 pandemic reported using hazard ratios and 95% confidence intervals (model adjusting for demographic factors only)**

|  | | **Hazard ratio** | **95% confidence interval** | **P value** |
| --- | --- | --- | --- | --- |
| **Cohort** | General population | Reference group |  |  |
|  | Clinically extremely vulnerable (CEV) children | 2·81 | 2.40–3.29 | <·001 |
|  | Children living with a CEV person | 1.09 | 0·97–1·22 | 0·14 |
| **Sex** | Male | Reference group |  |  |
|  | Female | 1·94 | 1·84–2.04 | <·001 |
| **Age group** | 2–7 | Reference group |  |  |
|  | 8–12 | 5.21 | 4·58–5·92 | <·001 |
|  | 13–17 | 19·39 | 17.20–21.86 | <·001 |
| **Deprivation quintile (WIMD 2019)** | 1 (most deprived) | 1·13 | 0·89–1·05 | <0.01 |
|  | 2 | 1.05 | 0·95–0.97 | 0·19 |
|  | 3 | 1·05 | 0·95–0.97 | 0·23 |
|  | 4 | 1·10 | 0·91–1·01 | <0.05 |
|  | 5 (least deprived) | Reference group |  |  |
| **Rural/Urban area** | Urban | Reference group |  |  |
|  | Rural | 1·02 | 0·98–0.97 | 0·45 |

**Multivariable analysis of risk factors for having a record of anxiety or depression during the COVID-19 pandemic reported using hazard ratios and 95% confidence intervals (model adjusting for both demographic and mental health factors)**

|  | | **Hazard ratio** | **95% confidence interval** | **P value** |
| --- | --- | --- | --- | --- |
| **Cohort** | General population | Reference group |  |  |
|  | Clinically extremely vulnerable (CEV) children | 2·27 | 1·94–2·66 | <·001 |
|  | Children living with a CEV person | 1·02 | 0·91–1·14 | 0·746 |
| **Sex** | Male | Reference group |  |  |
|  | Female | 1·58 | 1·50–1·66 | <·001 |
| **Age group** | 2–7 | Reference group |  |  |
|  | 8–12 | 4·56 | 4·01–5·18 | <·001 |
|  | 13–17 | 11·05 | 9·78–12·49 | <·001 |
| **Deprivation quintile (WIMD 2019)** | 1 (most deprived) | 1·02 | 0·95–1·10 | 0·565 |
|  | 2 | 0·98 | 0·90–1·06 | 0·614 |
|  | 3 | 1·00 | 0·92–1·09 | 0·965 |
|  | 4 | 1·07 | 0·98–1·16 | 0·127 |
|  | 5 (least deprived) | Reference group |  |  |
| **Rural/Urban area** | Urban | Reference group |  |  |
|  | Rural | 1·02 | 0·96–1·02 | 0·464 |
| **History of anxiety or depression** | No history | Reference group |  |  |
|  | Past history only | 5·13 | 4·75–5·53 | <·001 |
|  | Recent history only | 8·75 | 8·12–9·44 | <·001 |
|  | Both recent and past history | 18·98 | 17·52–20·55 | <·001 |

**Multivariable analysis of risk factors for having a record of anxiety or depression during the follow-up period March 23^rd^ 2019–January 31^st^ 2020 reported using hazard ratios and 95% confidence intervals (model adjusting for demographic factors only)**

|  | | **Hazard ratio** | **95% confidence interval** | **P value** |
| --- | --- | --- | --- | --- |
| **Cohort** | General population | Reference group |  |  |
|  | Clinically extremely vulnerable (CEV) children | 2·03 | 1.37–3.01 | <·001 |
| **Sex** | Male | Reference group |  |  |
|  | Female | 1·85 | 1·77–1·93 | <·001 |
| **Age group** | 2–7 | Reference group |  |  |
|  | 8–12 | 4·60 | 4.17–5.08 | <·001 |
|  | 13–17 | 18·50 | 16.89–20.27 | <·001 |
| **Deprivation quintile (WIMD 2019)** | 1 (most deprived) | 1·32 | 1.25–1·41 | <·001 |
|  | 2 | 1.22 | 1.15–1·31 | <·001 |
|  | 3 | 1·13 | 1.06–1·22 | <·001 |
|  | 4 | 1·09 | 1.02–1·17 | <0.05 |
|  | 5 (least deprived) | Reference group |  |  |
| **Rural/Urban area** | Urban | Reference group |  |  |
|  | Rural | 0.97 | 0·93–1·02 | 0·21 |

**Multivariable analysis of risk factors for having a record of anxiety or depression during the follow-up period March 23^rd^ 2019–January 31^st^ 2020 reported using hazard ratios and 95% confidence intervals (model adjusting for both demographic and mental health factors)**

|  | | **Hazard ratio** | **95% confidence interval** | **P value** |
| --- | --- | --- | --- | --- |
| **Cohort** | General population | Reference group |  |  |
|  | Clinically extremely vulnerable (CEV) children | 2·03 | 1.37–3.01 | <·001 |
| **Sex** | Male | Reference group |  |  |
|  | Female | 1·54 | 1·48–1·61 | <·001 |
| **Age group** | 2–7 | Reference group |  |  |
|  | 8–12 | 4·13 | 3.74–4.56 | <·001 |
|  | 13–17 | 11·56 | 10.53–12.68 | <·001 |
| **Deprivation quintile (WIMD 2019)** | 1 (most deprived) | 1·23 | 1.15–1·30 | <·001 |
|  | 2 | 1.14 | 1.07–1·22 | <·001 |
|  | 3 | 1·09 | 1.02–1·16 | <·05 |
|  | 4 | 1·05 | 0·98–1·13 | 0·17 |
|  | 5 (least deprived) | Reference group |  |  |
| **Rural/Urban area** | Urban | Reference group |  |  |
|  | Rural | 0.97 | 0·92–1·02 | 0·19 |
| **History of anxiety or depression** | No history | Reference group |  |  |
|  | Past history only | 4.50 | 4·21–4.81 | <·001 |
|  | Recent history only | 8·44 | 7.94–8.96 | <·001 |
|  | Both recent and past history | 16.97 | 15.85–18.18 | <·001 |
